# Investigation of Association Between microRNA Variants and Risk of Rheumatoid Arthritis

**DOI:** 10.1101/2022.07.29.22278172

**Authors:** Irfan Ullah, Muhammad Haseeb Tariq, Aftab Ali Shah, Muhammad Uzair, Mohsin Nawaz

## Abstract

Rheumatoid arthritis (RA) is a chronic autoimmune disorder that affects and damages the joints of human beings. It causes swelling, discomfort, and inflammation in and around the joints and affects other body organs; that affects 1% of the world’s population, with 6 to 60 people out of 100,000 developing the disease each year. However, a recent study describes that there are several factors involved in the regulation of RA disease, including genetic factors like MicroRNAs (miRNAs) variants, which are tiny molecules that bind to complementary target RNA molecules to regulate the protein-coding region of the genome, these non-coding short RNA molecules attach to target mRNA molecules at 3′-untranslated regions (UTR). The current study aimed to investigate the impact of variants rs11614913, rs6505162, and rs3746444 located in MIR196A2, MIR423, and MIR499, respectively, in RA patients. These SNPs were genotyped in RA patients and age- and gender-matched healthy controls using allele-specific T-ARMS-PCR. Allelic and genotypic frequencies of each variant were noted. Furthermore, from the selected variants, the association of rs11614913 and rs6505162 variants with the risk of RA was measured using a statistically odds ratio and confidence of interval (95%). In co-dominant models, the genotypic frequency of MIR423 variant rs6505162 was in cases A/A 75(35.21%), C/C 108 (50.7%) and A/C 30 (14.08%) while in controls A/A 34(16%), C/C 94(44.1%) and A/C 85(40%). These values indicate the best relationship between C/C and A/C, which were higher in cases and found significant association according to P-value and χ2. [χ2=14.03; P value=0.0009]. This concludes that SNPs (rs6505162) in MIR423 are the susceptibility factors for RA in the Pakistani Population. While SNPs rs11614913 in MIR196A2 have shown no association with RA.

## Introduction

Rheumatoid arthritis (RA) is a chronic inflammatory disorder characterized by severe inflammation in the joints. RA is marked by synovial inflammation and articular cartilage destruction, which leads to joint deformity [1]. In 2010, RA claimed the lives of around 48,000 people worldwide. Although the exact cause of RA is unknown, several environmental and genetic variables play a crucial influence in its progression [2].

In many auto-inflammatory disorders, the immune system plays a key role. Immune cells assault healthy joint tissues in RA, causing severe synovial inflammation [3]. he RA synovium’s large cell population primarily results from invasive and native cells’ deficient apoptosis [4]. Genome-wide association studies (GWAS) have discovered several hundred RA-associated variants. However, these recorded genetic variants only account for around 40%of the RA inheritance pattern [5]. GWAS data showed that the non-coding regions of the genome contain about 90% of disease-associated variants. This reveals the vital role of regulatory elements in the etiology of RA. Among these elements, MiRNAs are important post-transcriptional controlling molecules that regulate protein-coding genes by inhibiting protein synthesis or degrading target miRNAs. Several studies have shown the association of functional SNPs (rs11614913, rs6505162, rs3746444) in MIR196A2, MIR423, and MIR499A, respectively, with RA in different ethnic groups. Therefore, it is important to screen RA patients for the contribution of selected MiRNAs and SNPs in RA [6]. Apart from HLA genes, genome-wide investigations have indicated that numerous additional genetic risk factors are linked to the development of RA. Through genome-wide association studies (GWAS), more than 100 loci outside of HLA and SE have been found, accounting for5% of RA-linked genes [7].

Knowing genetic causes and the changes in amino acid sequence that they cause will help determine disease incidence and seriousness, which may lead to more tailored medicine. Furthermore, finding the biochemical and immunological pathways that define pathogenesis, which can then be used as a therapeutic target, is also significant [9]. Therefore, the present study was designed to investigate, in-silico, the coding sequence variants of the MIR196a2 and MIR423 genes. In addition, the current study also aimed to investigate the association between microRNA variants and with risk of Rheumatoid Arthritis.

## Material and Methods

Four hundred twenty-six people were chosen for this study (213 cases and 213 controls). Patients with RF levels greater than 15 u/ml and ACPA levels greater than 20μ/ml were declared positive and selected for blood collection. For DNA extraction, the Phenol-Chloroform extraction technique was used. Different chemicals used for DNA extraction are shown in Table 1**Error! Reference source not found**.

**Table 1.** Composition of different chemicals used in DNA extraction

For DNA confirmation and statistical analysis, the Agarose Gel electrophoresis (1.5%) and Nanodrop were used to measure the results (ThermoFisher Nano Drop 2000, Scientific). Allele-specific T-ARMS PCR was performed to detect specific SNP in target samples. PCR reactions were made in the specific proportion which is described in Table 2

**Table 2.** Composition of PCR mixture

| Chemicals | Composition |
| --- | --- |
| DNTPs | 1.0 µL |
| Mgcl<sub>2</sub> | 2.5 µL |
| 10x Taq Buffer | 2.5 µL |
| Forward Primer | 0.5 µL |
| Reverse Primer | 0.5µL |
| Taq DNA Polymerase | 1.0 µL |
| Template | 1.0 µL |
| Sterilized distilled water | 16 µL |
| Total Volume | 25 µL |

### Primers Designing

The studied genes MIR196A2 and MIR423 sequences were retrieved from dbSNP, and for our particular region, primers were designed using software primer 3. The details of primer sequences and their base pair lengths are listed in Table 3.

**Table 3.**
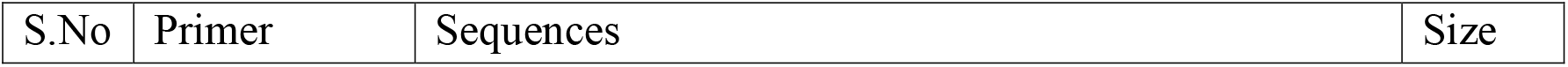
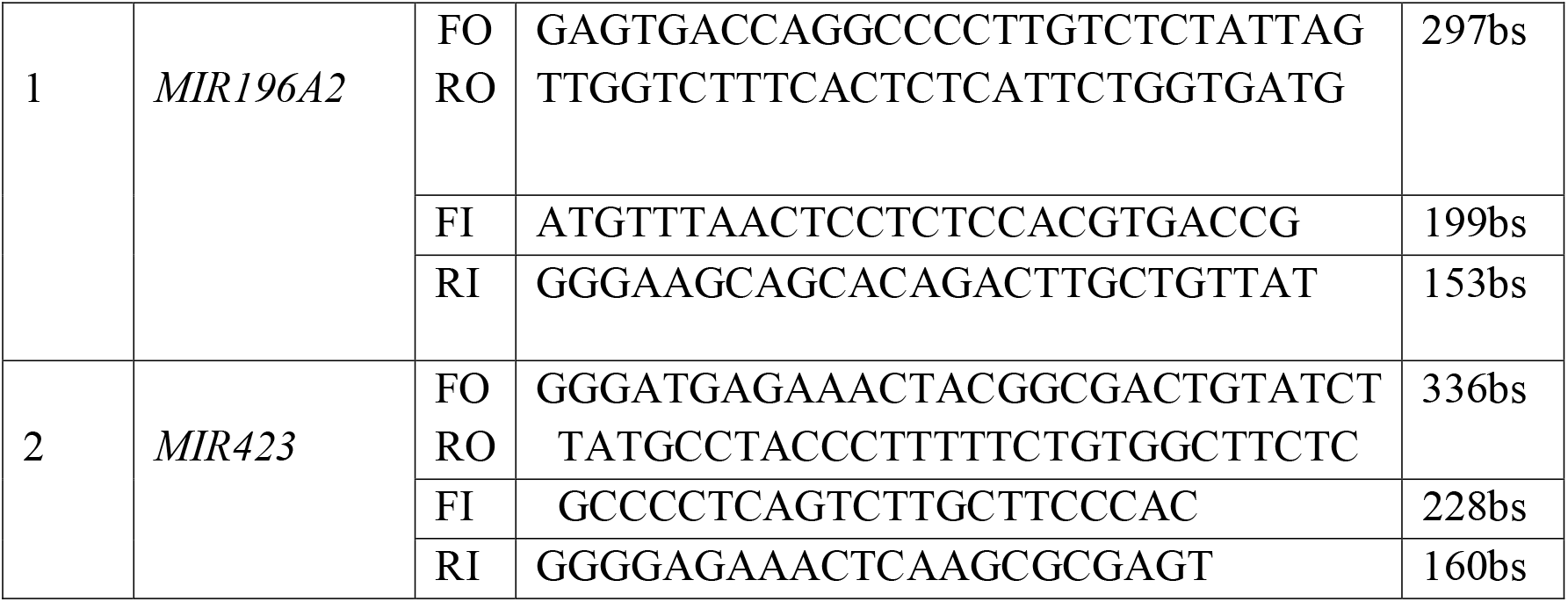
Primers for MIR196A2 and MIR423 and their sequences

**Table 4.** Genotypic distribution of variants rs11614913 in healthy and diseased samples

| Study subject | Genotype |  |  | X<sup>2</sup> Statistics |  |
| --- | --- | --- | --- | --- | --- |
|  | TT% | CC% | TC% | X<sup>2</sup> | P-value |
| Cases (n=213) | 93(44%) | 54(25.3%) | 66(31%) | 2.902 | P<0.23 |
| Controls (n=213) | 111(52.11%) | 30(14.08%) | 72(34%) |  |  |

**Table 5.**
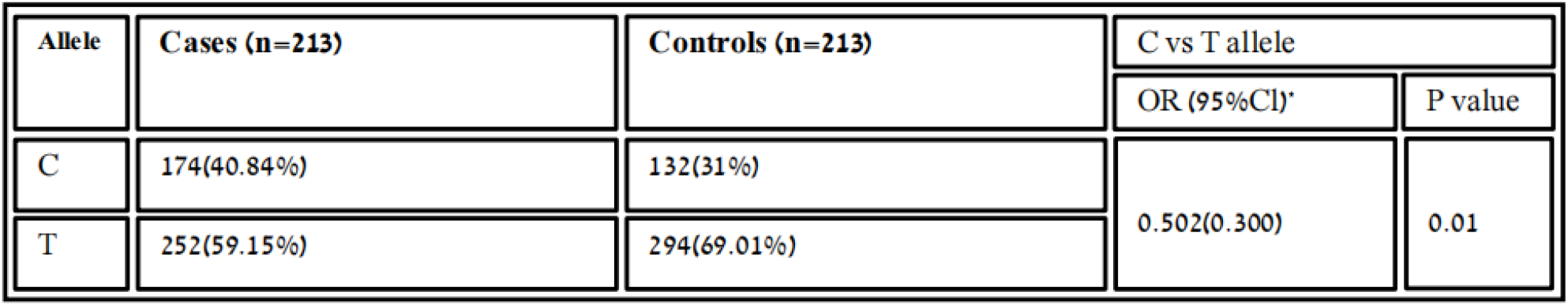
Distribution of allelic frequency of variant rs11614913 in healthy and patients samples

| Allele | Cases (n=213) | Controls (n=213) | C vs T allele |  |
| --- | --- | --- | --- | --- |
|  |  |  | OR (95%CI)* | P value |
| C | 174(40.84%) | 132(31%) | 0.502(0.300) | 0.01 |
| T | 252(59.15%) | 294(69.01%) |  |  |

**Table 6.** Homozygous dominant model association of variant rs11614913 between cases and control samples

| Model | Cases (n=213) | Controls (n=213) | TT vs TC+CC allele |  |
| --- | --- | --- | --- | --- |
|  |  |  | OR (95%CI)* | P value |
| TT | 93(51%) | 111(52.11%) | 0.949(0.478) | 1.00 |
| TC+CC | 90(51.1%) | 102(48%) |  |  |

**Table 7.** Homozygous recessive model association of variant rs11614913 between healthy and diseased samples

| Model | Cases (n=213) | Controls (n=213) | CC vs TC+TT allele |  |
| --- | --- | --- | --- | --- |
|  |  |  | OR (95%CI)* | P value |
| CC | 54(25.35%) | 30(14.08%) | 1.15(0.476%) | 0.82 |
| TC+TT | 159(74.64%) | 183(85.9%) |  |  |

**Table 8.**
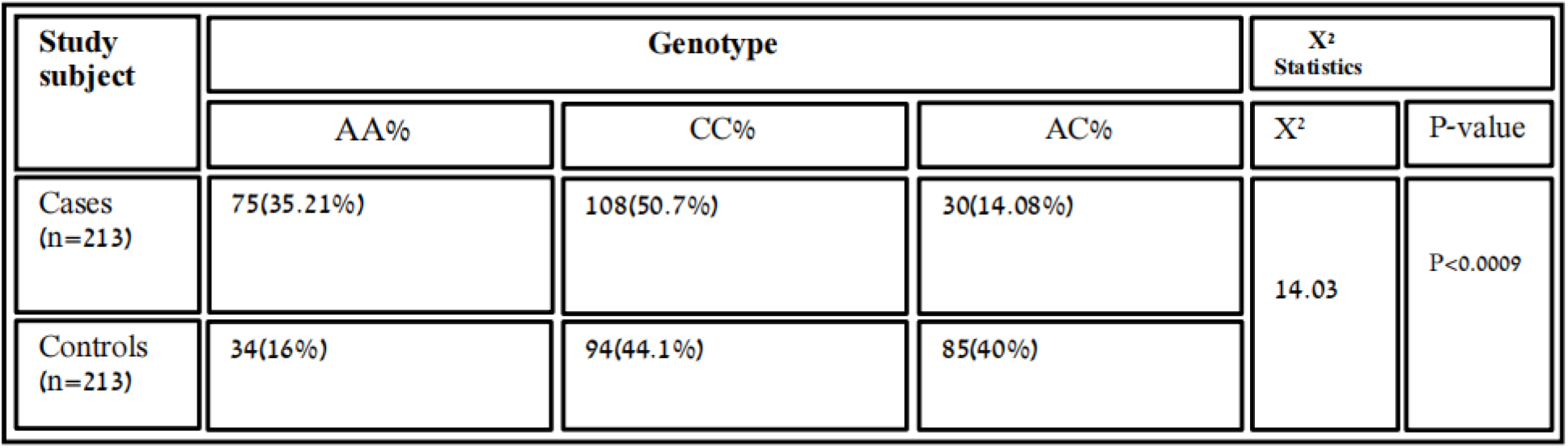
Association of variant rs6505162 with RA in cases and control samples

| Study subject | Genotype |  |  | X<sup>2</sup> Statistics |  |
| --- | --- | --- | --- | --- | --- |
|  | AA% | CC% | AC% | X<sup>2</sup> | P-value |
| Cases (n=213) | 75(35.21%) | 108(50.7%) | 30(14.08%) | 14.03 | P<0.0009 |
| Controls (n=213) | 34(16%) | 94(44.1%) | 85(40%) |  |  |

**Table 9.**
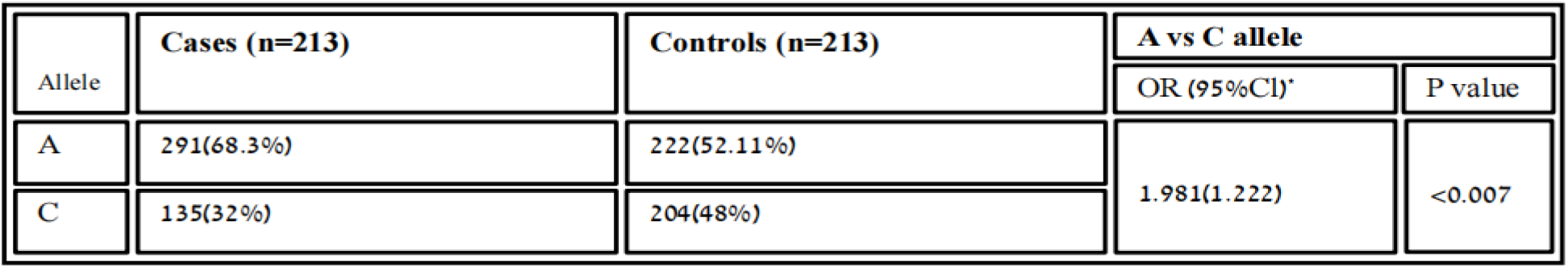
Distribution of Allelic frequency of variant rs6505162 in cases and healthy samples

| Allele | Cases (n=213) | Controls (n=213) | A vs C allele |  |
| --- | --- | --- | --- | --- |
|  |  |  | OR (95%CI)* | P value |
| A | 291(68.3%) | 222(52.11%) | 1.981(1.222) | <0.007 |
| C | 135(32%) | 204(48%) |  |  |

**Table 10.** Homozygous dominant model of the association between variant rs6505162 with RA in cases and controls

| Model | Cases (n=213) | Controls (n=213) | CC vs AA+AC |  |
| --- | --- | --- | --- | --- |
|  |  |  | OR (95%CI)* | P value |
| CC | 108(23.2%) | 96(50.70%) | 11.254(0.648) | 0.61 |
| AA+<br>AC | 105(67.5%) | 117(49.3%) |  |  |

**Table 11.**
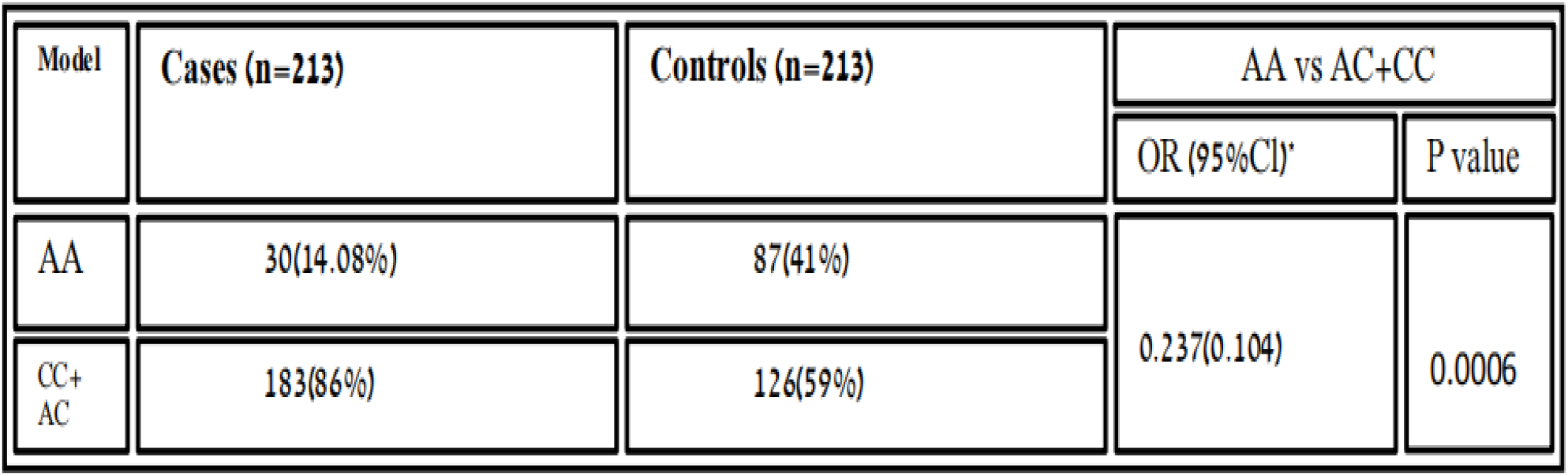
Homozygous recessive model of association variant rs6505162 with RA in healthy sample and cases

| Model | Cases (n=213) | Controls (n=213) | AA vs AC+CC |  |
| --- | --- | --- | --- | --- |
|  |  |  | OR (95%CI)* | P value |
| AA | 30(14.08%) | 87(41%) | 0.237(0.104) | 0.0006 |
| CC+<br>AC | 183(86%) | 126(59%) |  |  |

### Genotypic study of *MIR196A2* rs11614913

Using Amplification Refractory Mutation System-Polymerase Chain Reaction (ARMS-PCR), The MIR196A2 gene variant rs11614913 was genotyped, using four primers for amplifying our target portion. (Two forward primers and two reverse primers in total).

*MIR196A2*-FO: GAGTGACCAGGCCCCTTGTCTCTATTAG

*MIR196A2*-FI: ATGTTTAACTCCTCTCCACGTGACCG

*MIR196A2*-RO: TTGGTCTTTCACTCTCATTCTGGTGATG

*MIR196A2*-RI: GGGAAGCAGCACAGACTTGCTGTTAT

### Genotypic study of *MIR423* rs6505162

Similar to the above gene genotyping, the MIR423 gene variant rs6505162 was also genotyped by Amplification Refractory Mutation System-Polymerase Chain Reaction (T-ARMS-PCR), having four primers to amplify the target gene. (Two forward primers and two reverse primers). *MIR423*-FO: GGGATGAGAAACTACGGCGACTGTATCT

*MIR423*-RO: GCCCCTCAGTCTTGCTTCCCAC

*MIR423*-FI: TATGCCTACCCTTTTTCTGTGGCTTCTC

*MIR423*-RI: GGGGAGAAACTCAAGCGCGAGT

### Gel Electrophoresis

A thermal cycler reaction was completed, followed by gel electrophoresis through which an amplified reaction was carried out for genotyping, and the result was analyzed.

### Statistical Analysis

Graph Pad Prism Software 6 V was used to analyze genotypic and allelic differences in cases and controls using chi-square statistics. The Odd Ratio with a 95% Confidence Interval (CI) was also examined. Furthermore, the influence of homozygosity on the disease was investigated using homozygous dominant and homozygous recessive models. A statistically significant P value of 0.005 was used.

## RESULTS

### Role of *MIR196A2* rs11614913 T/C and *MIR423* rs6505162 C/A in Rheumatoid Arthritis

The association of genotypic and allelic frequencies for the SNP variants rs6505162 A/C and rs11614913 C/T in both healthy and controls are shown in Table 1 and Table 2. ll the genetic models were applied, such as dominant, co-dominant, recessive, and additive, on each variant and statistically tested as the association of MIR196A2 rs11614913 is given in (Table 1). Likely, the allelic frequency statistical analysis defined that there were significant changes in allele T and C frequency in both groups (OR 0.502(0.300-0.840) P<0.01 (Table 1, Figure 2). The frequency of allele T was higher in the control sample was 294 (69.01%) as compared to cases 252 (59.2%), whereas allele C frequency was 174 (40.84%) in diseased and 132 (31.6%) in healthy samples. For the association of variants rs11614913 with Rheumatoid arthritis, both homozygous models, dominant (CC vs. TT+TC) and recessive (CC vs. TC+TT), were used (Table 1) to confirm genotyping errors, 10% of the total samples were checked again.

**Figure 1.**
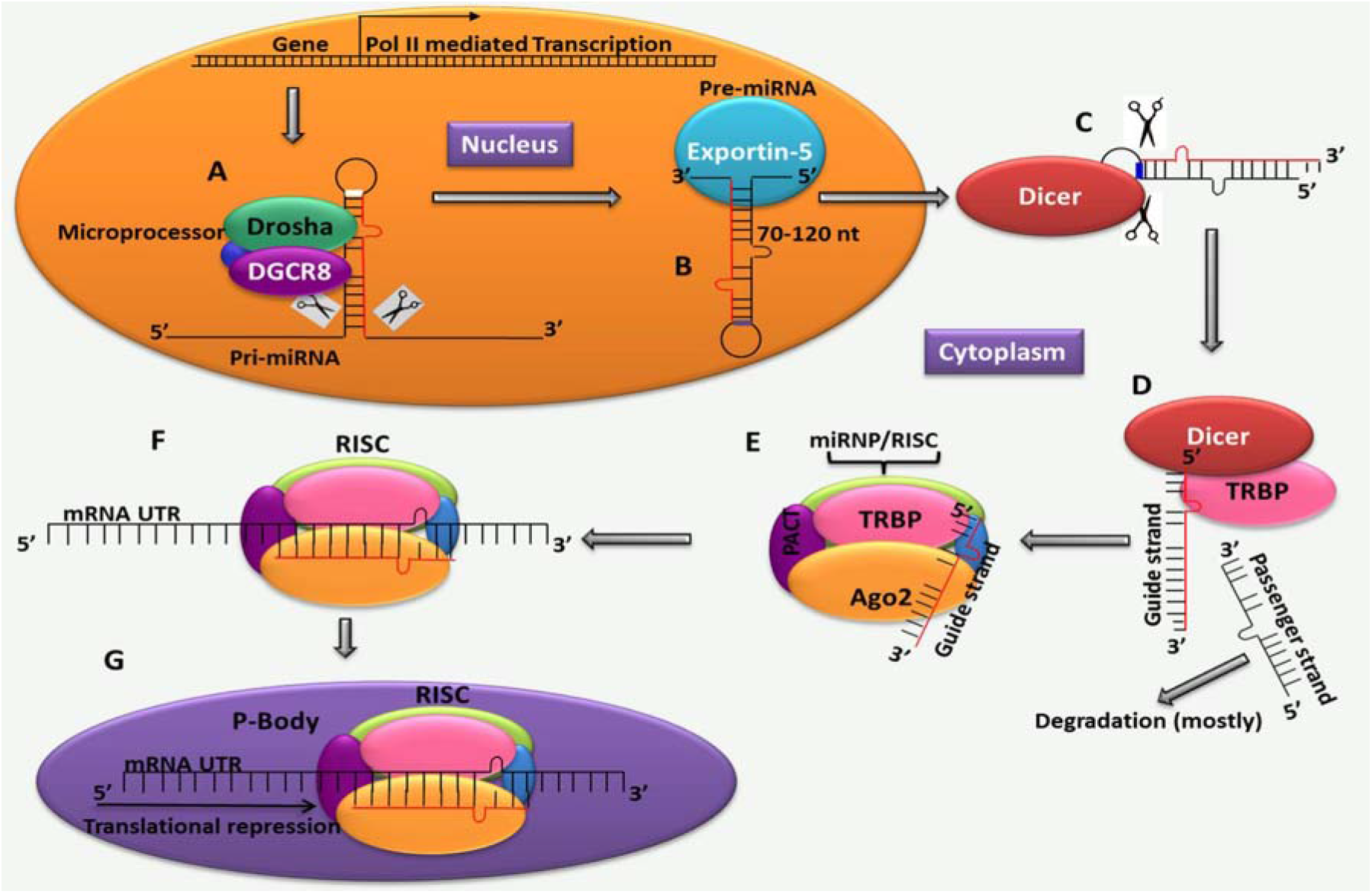
Synthesis of MiRNAs and their modification [8]

**Figure 2.**
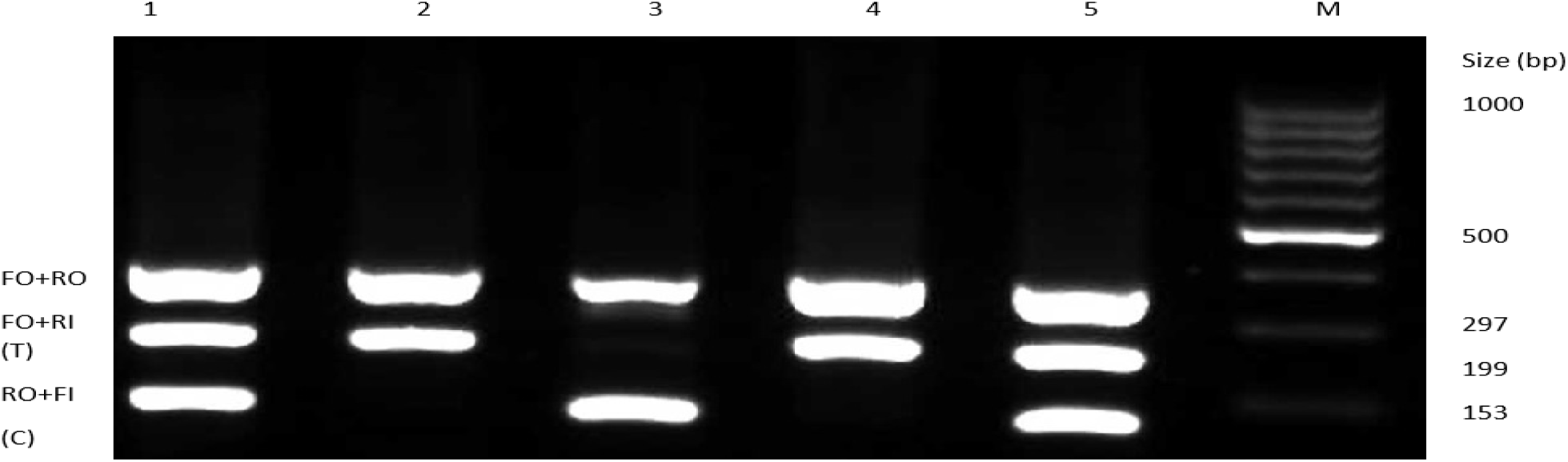
Representative Gel picture of the genotypic distribution of variants rs11614913 of healthy and diseased samples,

In Figure 3, the uppermost lane, FO+RO, represents the outer region of the gene, which is 297 bp in size. The FO+Ri strands are T-represented strands about the size of 199 bp, and the RO+Fi represents allele C having a size of 153 bp.

**Figure 3.**
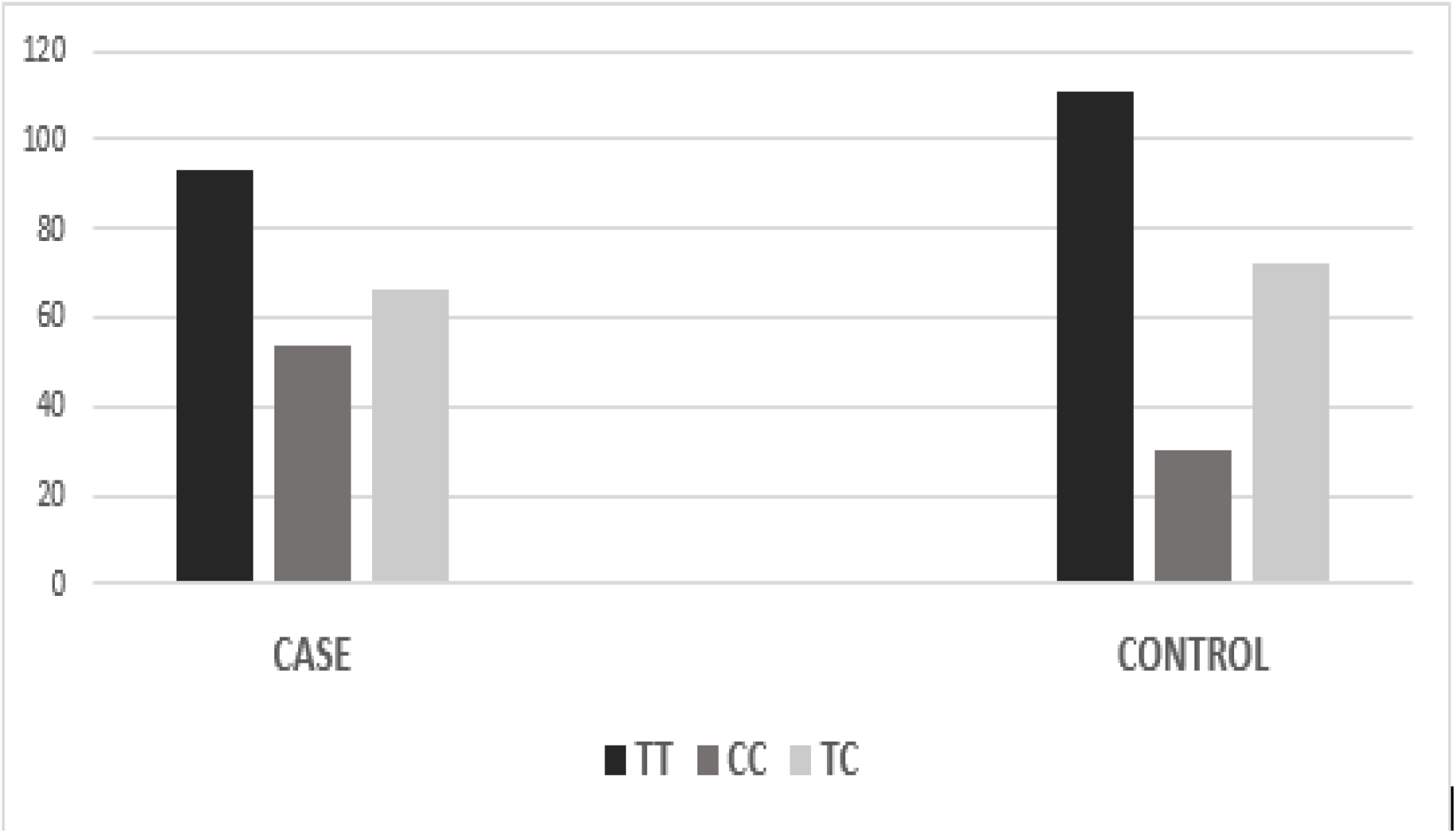
Graphical analysis of the genotypic distribution of variant rs11614913

### Association of variant rs6505162 with rheumatoid arthritis in healthy and diseased samples

According to co-dominant models, the genotypic frequency of MIR423 rs6505162 observed in cases was A/A 75(35.21%), C/C 108(50.7%), and A/C 30 (14.08%), while in controls, A/A 34 (16%), C/C 94(44.1%) and A/C 85 (40%) and found significant association [χ2=14.03; P value=0.0009]. whereas, the allelic distribution showed a higher frequency of allele A 291(68.3%) in cases while in control 222(52.11%) expression compared to allele C, which was 135 (32%) in cases and 204(48%) in controls.

In *Figure 5*, the 1st and 2nd well have a heterozygous condition of AC, the 3rd well has a homozygous condition of AA, and the 4th well has a homozygous CC allele.

**Figure 4.**
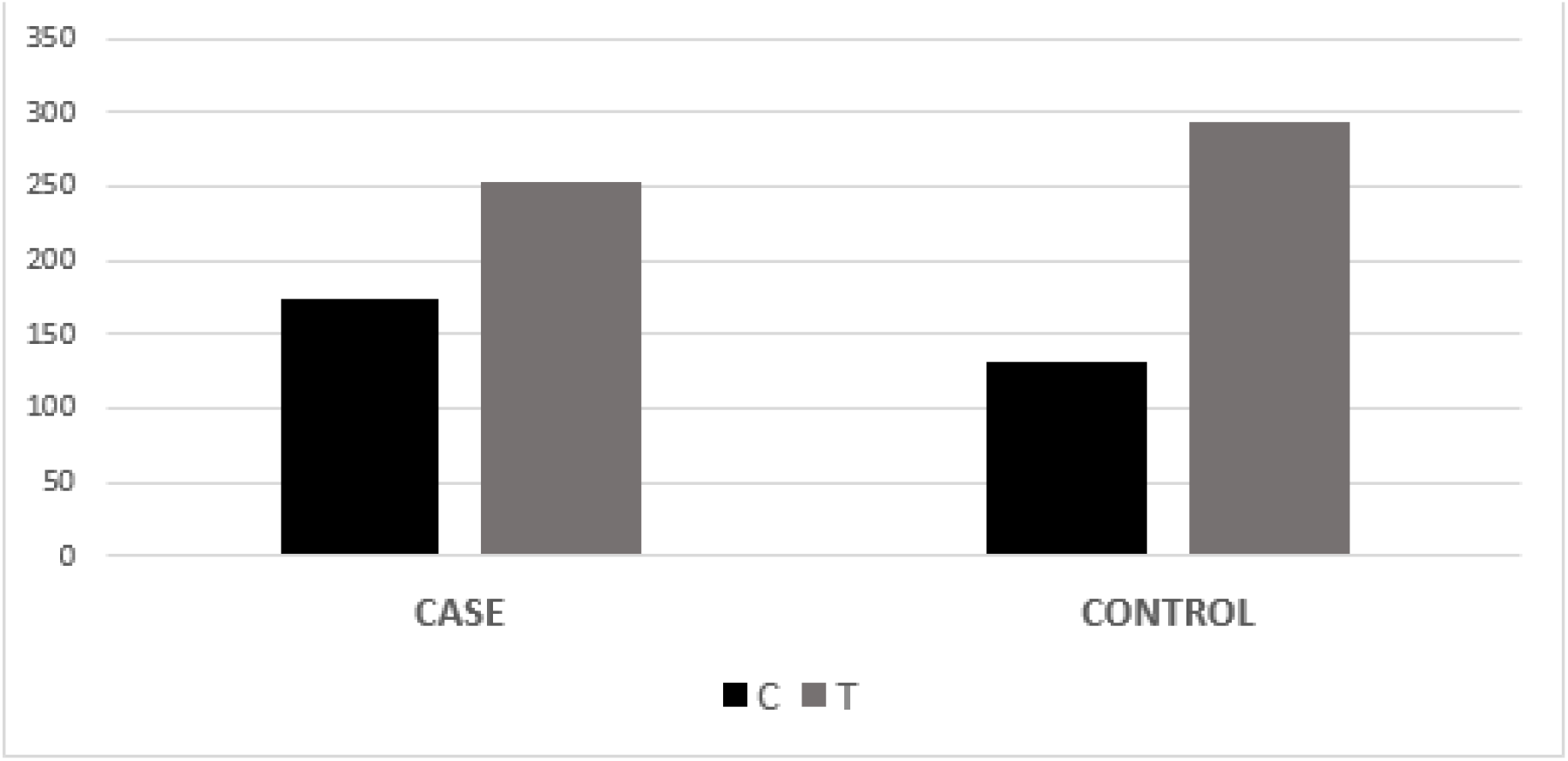
Graphical analysis of the allelic frequency of variant rs11614913 between cases and control samples

**Figure 5.**
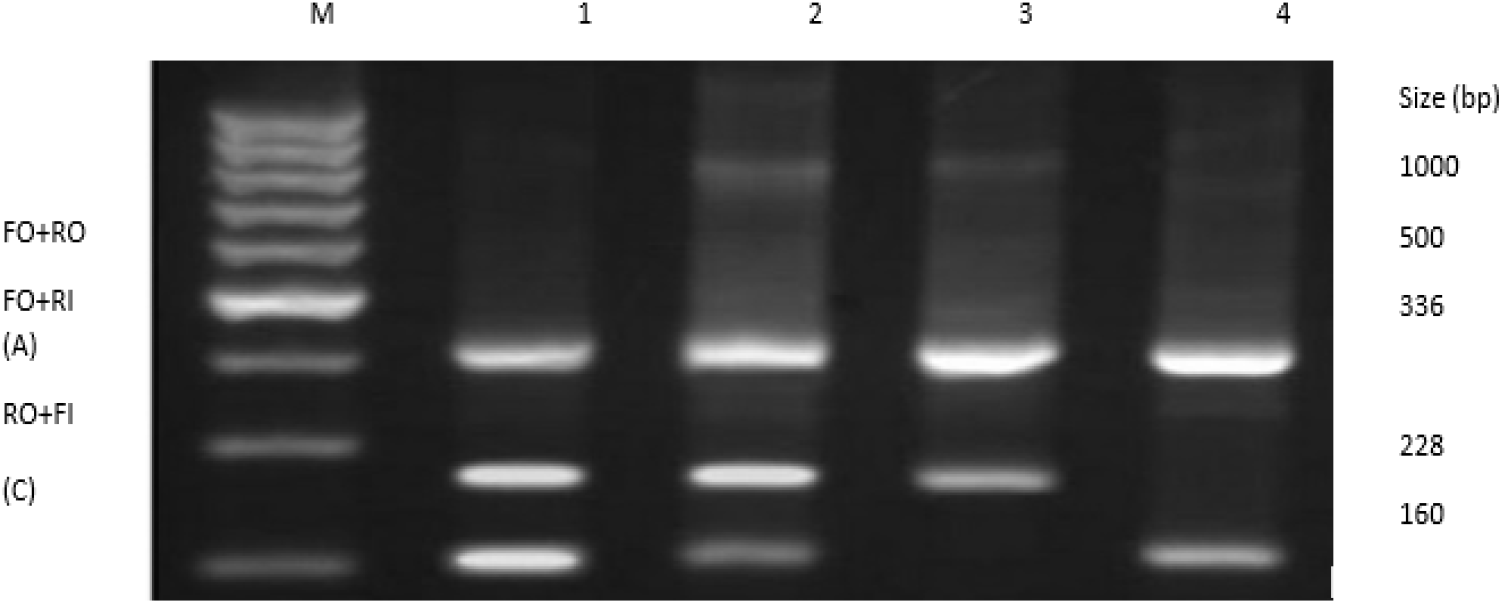
Gel picture genotypic distribution of variant rs6505162 in cases and controls samples

**Figure 6.**
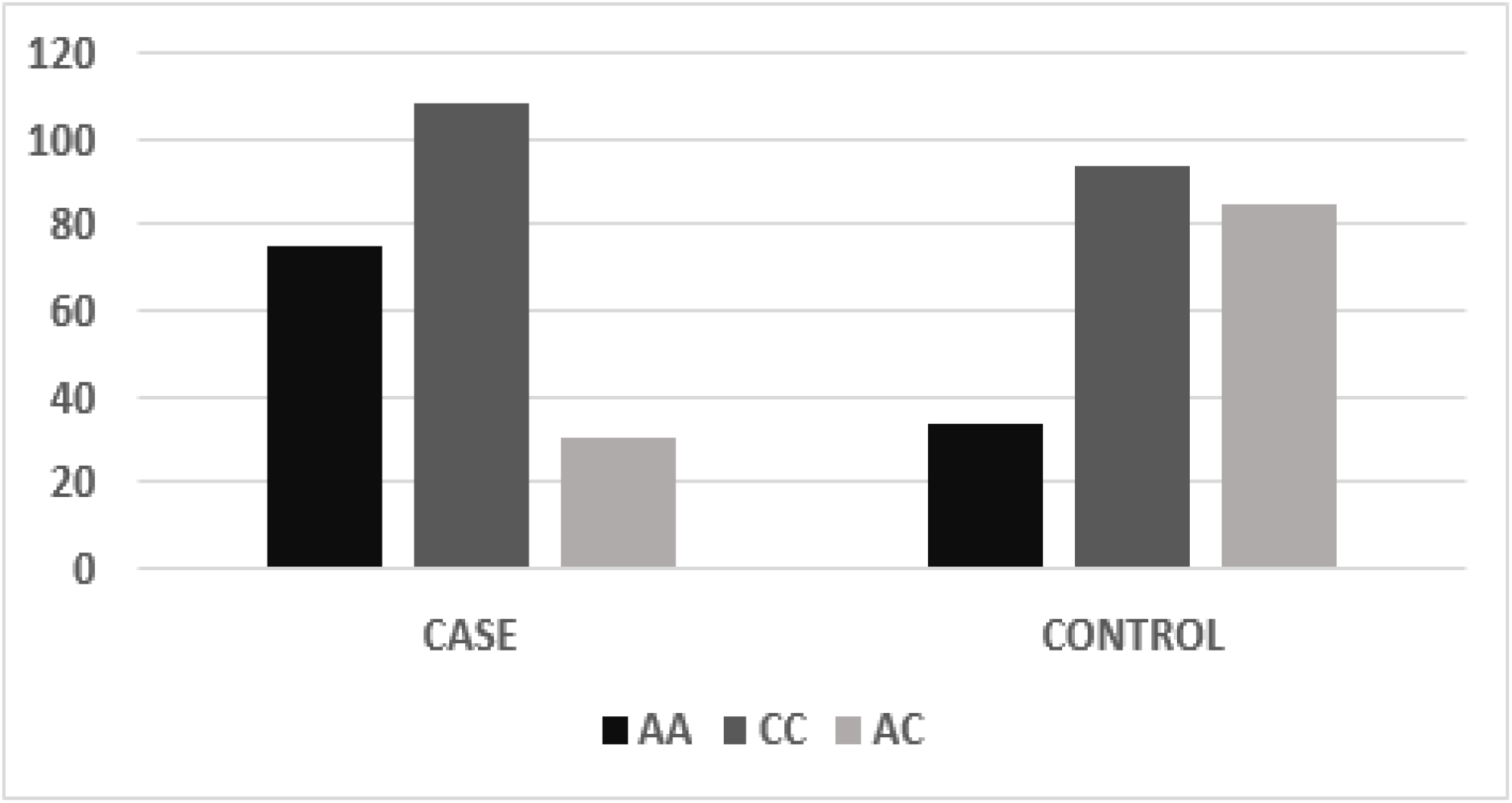
Graphical presentation of genotypic frequency of MIR423 rs6505162 in cases and controls

**Figure 7.**
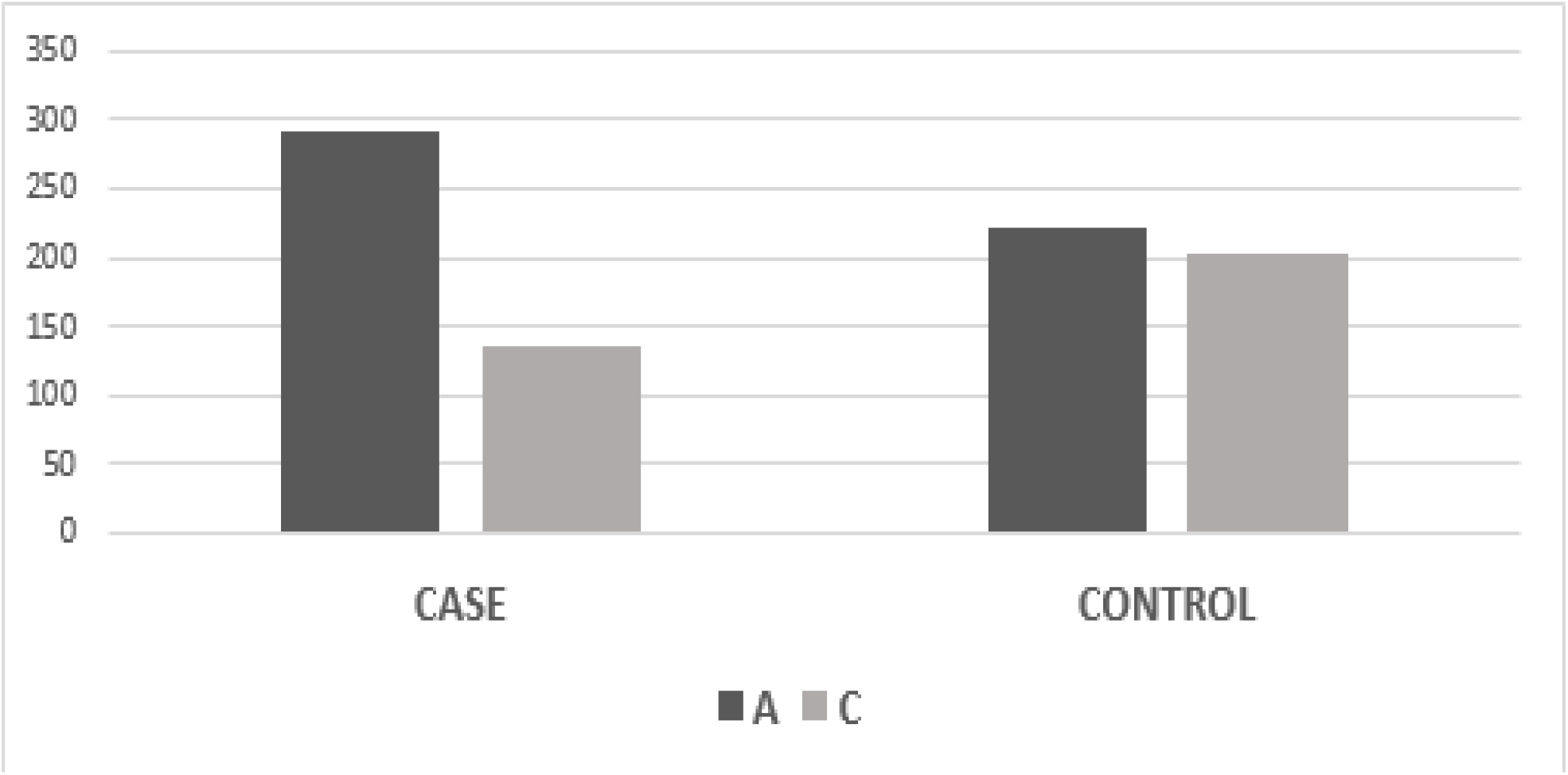
Graphical presentation of allelic frequency of *MIR*423 rs6505162 in Cases and controls

**Figure 8.**
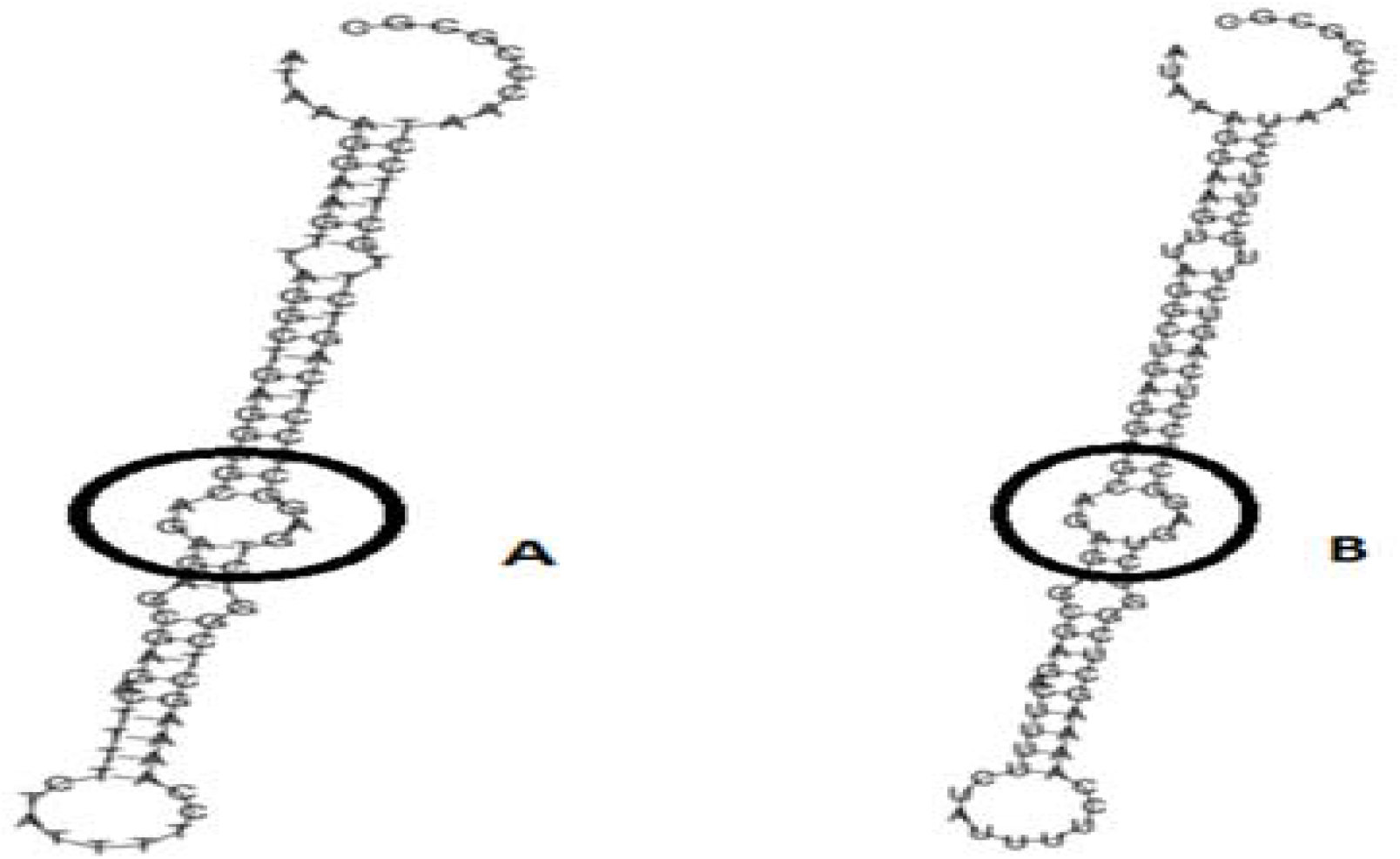
*MIR423* secondary structure prediction show the lower and higher energy state due polymorphism in its structure

**Figure 9.**
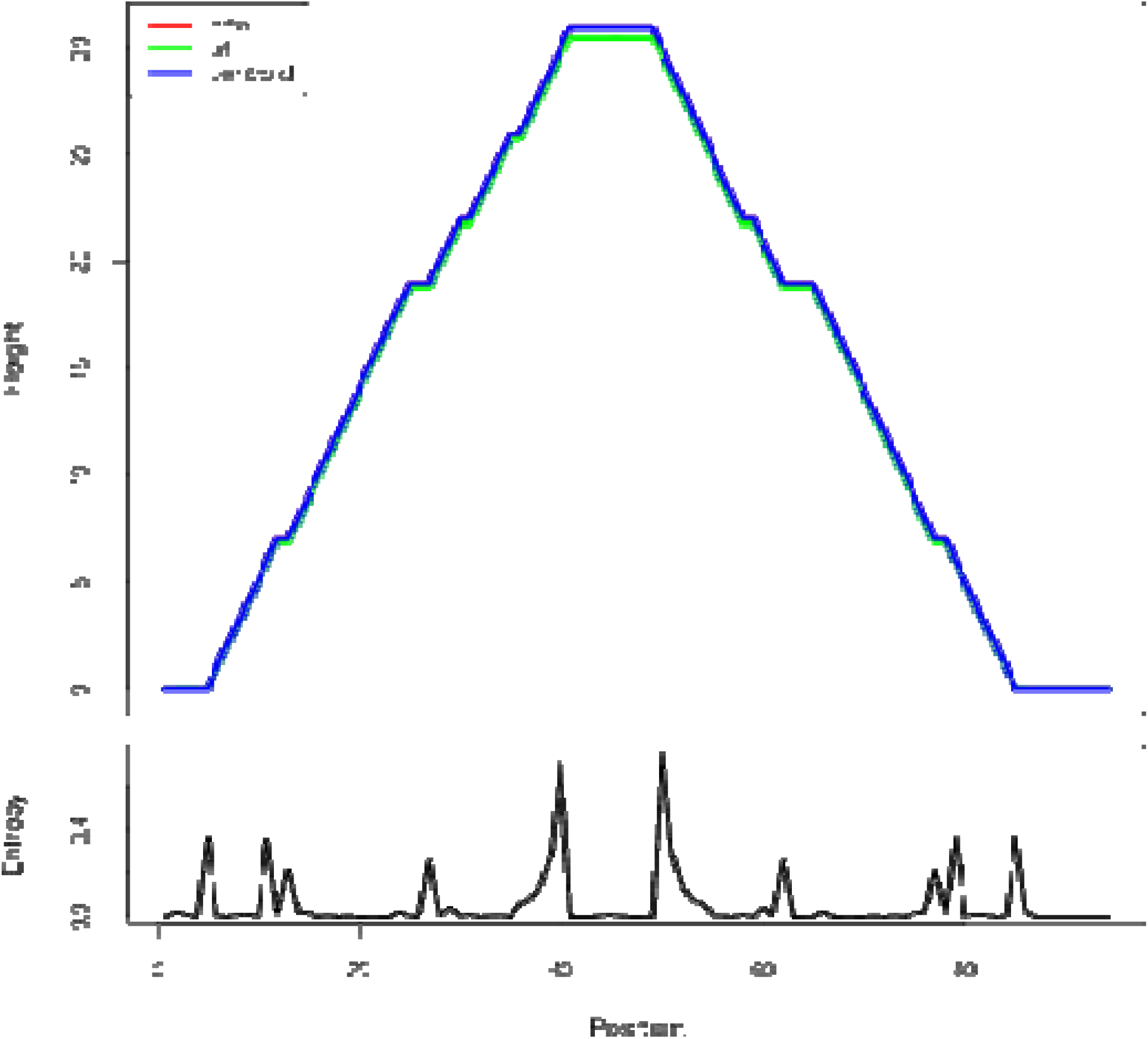
Mountain plot presentation of MIR423 secondary structure prediction

### Secondary Structure Prediction

MIR423 secondary Structure prediction base-pair coding probabilities with minimum free energy is (48.80 kcal/mol Allele T), and encoding MFE base-pair probabilities with minimum free energy (49.90; Allele U), and the MFE structure frequency in the ensemble is 62.63 %. For the presentation variant MIR423 alleles, a mountain plots graphical assessment of the minimal free energy diagram, the RNA thermodynamic ensemble structures, and the centroid structure was created. The stabilized lower energy level of MIR423 has the band between base A with base T that is shown in the below encircle diagram, while in a higher energy state, the base T is replaced by base U, which causes the regulation, the structure of MiRNA as well as in the function of it.

## Discussion

The current study shows that the MIR423 is vital in regulating Rheumatoid Arthritis in the Pakistani population. Using different perspectives, such as family linkage and genome-wide association studies, scientists can discover the genetic risk factors in many monogenic and polygenic diseases (GWAS). Because RA is such a complicated illness, scientists have been trying to determine genetic risk factors. As we know, immune system regulation is essential in preventing a variety of pathogenic disorders, such as autoimmune disease and cancer, for that mammal has evolved a complex system of checks and balances for immune regulation to maintain self-tolerance while allowing immune responses to foreign pathogens, the majority of which are unknown. MiRNAs have recently been discovered to have a crucial function in controlling immune response and immune cell growth. Yet, only a few particular MiRNAs have been identified as significant immune system regulators [10]. The etiology of RA is diverse, including many cell types associated with the innate and adaptive immune systems. RA begins as a chronic state of cellular activity that leads to autoimmunity and immune complexes in the joints and other organs where it appears [11]. Genetic and epigenetic factors play key roles in autoimmune diseases. For the regulation of leukocyte activation and cytokine production, MiRNAs are important, which result in autoimmune disease [12]. MicroRNAs (MiRNAs) are short non-coding RNA molecules of 19–23 nucleotides in length. They suppress the expression of numerous protein-coding genes via increasing messenger RNA (mRNA) degradation or translational repression at the post-transcriptional stage. MiRNAs have a role in various functions, including cell proliferation, differentiation, inflammation, and signal transduction. MiRNAs have a role in developing various illnesses, including cancer, diabetes, and heart disease.

Changes in miRNA status and their targets may help researchers figure out which pathways are involved in the etiopathogenesis of autoimmune diseases. MiRNAs have been suggested as potential biomarkers for both the diagnosis and prognosis of autoimmune disorders In autoimmune diseases, MIR196A2 and MIR423 have received a lot of attention. The single base mutation in rs11614913 found at 3arm (3p) of MIR196a2 mature sequences was believed to dysregulate its full function and secondary structure that may affect the expressions of its target messenger RNA. Annexin A1 (ANXA1) regulation is also directed by rs11614913, which maintains the expression and regulation of cyclooxygenase 2, phospholipase A2 and nitric oxide synthase. The change T from C accelerates the expression level of the MIR196A2 gene, adversely affecting the expression of TNF-α, which works in inflammation and thrombosis [14]. MIR423 regulates such PA2G4 expression.

PA2G4 protein levels have been observed in MIR423 mimic-or MIR423 inhibitor-treated HEC-1b cells. A research study of the Iranian population described pre-MIR196A2 variant rs11614913 change as a potential causing factor of rheumatoid arthritis (RA) and common lupus (SLE). The above study described a significant increase in RA patients carrying the T Vs. The c allele of the pre-MIR196A2 variant rs11614913 and the genotypic association of TC of the pre-MIR196A2 variant rs11614913 greatly reduced the chance of RA [15]. Another study was conducted in the Egyptian population to report the genetic polymorphisms of MIR196a2 rs11614913 to analyze susceptibility to RA. The valuable changes were studied in classifying the snRNPs genotypes and their minor alleles in RA patients and healthy samples. The prevalence of MIR196A2 variant rs11614913 and their C alleles significantly increased in RA patients.

Furthermore, factor X4 production is also regulated by the modification of MIR196A2. That affects DRB1 antigen regulation, which is associated with the risk of RA Also, AMI patients have been studied to show an increased amount of MIR423-5p. Interestingly, the amount of MIR423-5p expression has increased in a subgroup of the same acute myocardial infarction patients six months after the initial condition. The selected MIR196A2 and MIR423 roles have been mentioned in different populations. The polymorphisms in MiRNA196A2 and MIR423 are associated with increased CAD risk in Korean populations. It is also investigated that MIR196A2 rs11614913 polymorphism results demonstrate significant associations with cancer and have a role in Systemic Lupus Erythematosus diseases [17].

In people who smoke, the Hsa-MIR423 (rs6505162) gene has been linked with esophageal squamous cell carcinoma elevated risk [18]. According to recent research, MIR423, rs6505162 also has the potential to become a biological marker for analyzing several disorders such as CAD [19]. In our results, the genotype frequency of MIR423 rs6505162 in RA was observed in cases A/A 75(35.21%), C/C 108(50.7%), and A/C 30(14.08%) while in controls A/A 34(16%), C/C 94(44.1%) and A/C 85(40%). These values indicate the best relationship between C/C and A/C, which were higher in cases and found significant association according to P-value and χ2. [χ2=14.03; P value=0.0009]. Whereas the allelic distribution shows a higher frequency of allele A 291(68.3%) in cases while in control 222(52.11%) expression compared to allele C, which is 135(32%) in cases and 204(48%) in controls. The effect of the homozygous dominant model of MIR423 showed higher TT 111(52.11) in controls and in cases 93(43.66%), while TC+CC were 102(48%) looked higher compared to cases which were 90(51.1%). And homozygous recessive model showed allele CC higher in cases 54(25.35%), and in controls, 30(14.08%), and TC+CC was 159(74.64%) in cases and 183 (85.9%). The MIR423 (rs6505162) indicates this miRNA might be associated with RA risk in the Pakistani population.

The miR-196a2 rsl11614913 C allele with coronary heart disease increased the risk of myocardial infarction and other serious cardiovascular events. And communities in southeastern China [20]. In our results, the genotype frequency of MIR196A2 rs11614913 in RA was observed in cases T/T 93(44%), C/C 54(25%) and T/C 66 (31%) while in controls T/T 111 (52.11%), C/C 30(14.08%) and T/C 72 (34%) and found insignificant association [χ2=2.902; P value=0.23]. In Pakistani populations, the allelic distribution showed a higher frequency of allele T 294(69%) in control, while in cases 252(59.11%) expression compared to allele C, which is 174 (40%) in cases and 132(31%) in controls. The effect of the homozygous dominant model showed higher TT 111(52.11) in controls and in cases 93(43.66%), while TC+CC were 102(48%) looked higher compared to cases which were 90(51.1%). And homozygous recessive model showed allele CC higher in cases 54(25.35%), and in controls, 30 (14.08%), and TC+CC was 159(74.64%) in cases and 183 (85.9%). The increased production of interleukin has been well established in RA, as it is one of the dangerous autoimmune illnesses. In Pakistani RA patients, we discovered a link between MIR423 rs6505162 and MIR196A2 rs11614913 and RA. We looked at the impact of several models on the relationship and found that they were all positive. Although we used a limited sample size, a larger study with a larger sample set is needed to substantiate this link. This study can aid physicians’ incorrect management and therapeutic intervention if used properly.

## Conclusions

The current study investigated the association of MIR423 variant rs6505162 and MIR196A2 variant rs11614913 with the risk of rheumatoid arthritis patients in Pakistan. It showed the best relationship and involvement of these miRNAs regulation in 213 RA patients, as the same amount of healthy samples were treated by TARMS-PCR. Interestingly, the MIR423 variant rs6505162 showed significant association in the regulation of RA, using the statistical measurement tools like Odd ratio and Confidence Interval along with a significant P value (0.005). Furthermore, the genotypic and allelic frequency was measured following the Hardy-Weinberg principle using all models, such as the dominant, recessive, co-dominant, and additive models. On the other hand, the MIR196A2 rs11614913 single base change was normal for both the healthy and diseased samples to show that the dysregulation of this MiRNA does not affect the regulation of Rheumatoid arthritis.

## Future Recommendation

Rheumatoid arthritis is a joint disorder in human beings, while the exact pathogenicity of RA is known and increasing daily. To overcome this issue, it is important to find out all the factors involved in the cause of RA. For one, it is believed that miRNA variants have played a role in regulating RA. Therefore, the current study was conducted to find the correlation between MIR423 variant rs6505162 and MIR196A2 variant rs11614913 with the risk of RA. It was a great achievement and addition of knowledge to the future study that the selected miRNAs showed a significant association and role in the regulation of rheumatoid arthritis in the Pakistani population.

In contrast, the limited area of population and the number of miRNAs were selected. It would be beneficial to increase the size of samples and the number of variants. The advanced bioinformatics tools and more specific types of PCR may help reach the etiology of RA; the earlier stage of RA compared to the late stage can easily be diagnosed and treated. Advanced tools like Alu PCR and Allele-specific PCR might be possible to investigate the cause of RA and the role of miRNAs.

## Data Availability

The findings of this study are available within the article.

